# Young people’s experiences of green social prescribing at a community farm and perceived influences on mental health and wellbeing: a qualitative study in Wales

**DOI:** 10.64898/2026.11.17.26368284

**Authors:** Alison J Wainwright, Menna Brown

## Abstract

Mental health difficulties often begin in adolescence and early adulthood, yet most evidence on green social prescribing, which connects people with nature-based community activities to support health and wellbeing, comes from adult and quantitative research. This qualitative study explored how young people experienced taking part in a community farm programme in Wales and how they felt it influenced their mental health and wellbeing. Twelve young people aged 16 to 25 years took part in individual, semi-structured interviews at Swansea Community Farm in 2026. Data were analysed using reflexive thematic analysis within an interpretivist paradigm.

Four interconnected themes were developed: ‘belonging in a supportive community’; ‘finding purpose through meaningful participation’; ‘personal growth and enhanced wellbeing’; and ‘nature as the context for wellbeing’. Participants rarely described nature alone as responsible for feeling better. Instead, the farm brought together people who accepted them, useful work with real responsibility, and time outdoors with animals, and these were experienced together rather than separately. Participants had found the farm informally, and none had been referred through a formal social prescribing pathway. These findings suggest that green social prescribing for young people may work best when it is designed around belonging, meaningful roles and responsibility, with nature as the setting that makes these possible, and that evaluation should capture belonging, confidence and purpose as well as symptom reduction. They also point to the need for clear, youth-friendly routes into community provision, including self-referral. This was a single-site study, so it cannot show causal effects or represent the experiences of young people who did not engage, but it offers an in-depth account to inform the design and evaluation of provision for young people.

## Introduction

Poor mental health among young people is one of the most pressing public health problems globally. Adolescence and early adulthood are important developmental periods for prevention and health promotion [1,2], and they are also when most mental health difficulties first appear. A global meta-analysis of 192 epidemiological studies estimated that around half of all mental disorders begin before the age of 18 and more than 60% before the age of 25 [3]. These are years of rapid change in education, work, relationships and independence, and it has been argued that adolescence is better understood as lasting into the mid-twenties [2]. Surveys suggest that around one in five children and young people in England has a probable mental health disorder [4], and demand has outpaced the capacity of specialist services, with rising thresholds for access to child and adolescent mental health services [5].

The consequences reach well beyond health services. Young people with mental health difficulties are more likely to be out of education, employment or training later on, a status that is itself associated with poorer mental health [6]. Across the whole population, mental ill health was estimated to cost England around £300 billion in 2022, with most of that borne by people living with mental health difficulties and their families through lost earnings, reduced quality of life and informal care [7]. Difficulties that start early can therefore carry costs for individuals, families, employers and public services for many years. This is one reason why approaches that help young people stay well, and that sit alongside rather than inside stretched clinical services, are attracting growing interest.

Social prescribing is one such approach. It connects people with activities and sources of support in their community, often with the help of a link worker, to address the social and practical factors that shape health [8,9]. In Wales, it sits within national policy, and the national framework recognises several routes into provision, including self-referral, referral from statutory services outside health, and referral from healthcare [10]. This builds on wider commitments to wellbeing and prevention [11–14]. Green social prescribing (GSP) links people with activities undertaken in natural environments, including community gardening, conservation, care farming and volunteering [8,9]. The same idea is often called nature-based social prescribing (NBSP) in the literature. We use GSP throughout.

The evidence for GSP is encouraging but uneven. Reviews, mostly of adults, report potential benefits for mental wellbeing [15,16], anxiety and depression [15,16], and social connectedness [17,18], while also describing wide variation in intervention design, populations and outcome measures [15,19]. Systematic reviews of social prescribing more broadly have repeatedly noted small studies, high risk of bias, a lack of comparison groups and short follow-up [20]. We also know little about which features of GSP matter. Qualitative studies with adults point to relationships with staff and peers, a sense of belonging, learning new skills and contact with nature itself [21–23]. These accounts suggest that GSP may work through social and occupational processes as much as through nature exposure, but they were gathered from adults.

Much less is known about young people. Social prescribing has largely been developed and evaluated with adults, and there is limited evidence about whether it is acceptable to young people or what they themselves want from it [24]. Research on nature and the mental health of children and young people suggests potential benefits, but the interventions studied are very diverse [25,26]. The small number of qualitative studies of young people in GSP-type activities, with young pregnant women [27], care leavers [28] and students in a campus garden [29], report social connection and the restorative qualities of nature as important, but each involves a particular group in a particular setting. Access is a further question. In a recent study of stakeholders in South Wales, one participant asked who was covering young people, given that social prescribers were thought to work mainly with adults [20].

Community and city farms offer a distinctive setting for GSP. These are typically charity-run working farms where people look after animals, grow food and maintain the site alongside others, and in doing so contribute to the running of a place that the local community uses and values. Care-farming research, mostly with adults, suggests benefits for quality of life, depression and anxiety, although the evidence base is small [30], and broader qualitative evidence suggests that social relationships and shared purpose may matter alongside contact with nature [15,31]. What is missing is an account from young people themselves of what a community farm offers them and which parts of it they find meaningful.

That gap matters for practical reasons. If GSP is going to be offered to young people, commissioners and providers need to know what to build into it, who it suits and why, and how young people will find their way to it. Qualitative research can help with each of these. It can show which features of provision young people value and would need protecting if schemes were scaled up or evaluated, and which outcomes matter to them but may be missed by standard symptom measures [19,21]. It can also show how young people reach provision in the first place, which bears directly on how referral pathways should be designed.

This study therefore explored young people’s experiences of taking part in a community farm programme in Wales and their perceptions of how taking part influenced their mental health and wellbeing.

## Materials and methods

### Ethics

Ethical approval was granted by the Swansea University, Faculty of Medicine, Health and Life Sciences, School of Health and Social Care, Research Ethics Committee (reference 2 2026 15441 16277; approved 5 May 2026). Written informed consent was obtained from all participants, including those aged 16–17 years, and the committee approved this approach.

### Study design and methodological approach

A qualitative design employing semi-structured individual interviews was used to explore subjective experiences and meaning. The study was situated within an interpretivist paradigm, which treats knowledge as constructed through people’s accounts of their own experience and through the researcher’s interpretation of those accounts [32,33]. The study is reported in line with the Consolidated Criteria for Reporting Qualitative Research (COREQ) [34], and the completed checklist is provided as S1 Checklist.

### Aim and objectives

The aim was to explore young people’s experiences of taking part in a community farm programme and their perceptions of how participation influenced their mental health and wellbeing. The objectives were to: (1) understand how young people found their way to the farm; (2) describe their experiences of the farm, including relationships with staff and other participants, the activities, and contact with nature and animals; (3) explore how they felt participation influenced their mental health and wellbeing, including any challenges; and (4) consider what the findings mean for the design of, and access to, green social prescribing for young people.

### Setting and participants

Swansea Community Farm is a city farm in South Wales (UK). The farm provides weekly volunteering and community-focused activities including animal care, horticulture, conservation, education and training for those in the local community. Its programmes aim to reduce social isolation and loneliness, address poor mental health and improve wellbeing. Young people take part in weekly sessions choosing animal husbandry or land-based activities such as gardening, woodwork or conservation.

### Recruitment and data collection

Participants were purposively sampled. Eligibility criteria were age 16–25 years, current volunteering at the farm, and capacity to provide informed consent. The farm operation manager acted as gatekeeper and informed eligible volunteers about the study during regular farm activities and morning briefings. Interested volunteers arranged participation independently; it was clearly explained that participation was voluntary and that there were no consequences for non-participation.

Interviews took place between June and July 2026, conducted by the first author (AW). Each participant completed one individual interview after written informed consent was obtained. Most interviews were held in a private room; one took place in an outdoor safari tent when the indoor room was unavailable. The interviews explored motivations for taking part, experiences of the farm, perceived influences on mental health and wellbeing, relationships with staff and other volunteers, experiences of nature and animals, and challenges encountered.

### Data analysis

Interviews were audio-recorded and transcribed using Microsoft Word’s transcription function, and the transcripts were manually checked (AW) against the original recordings. Transcripts were anonymised. Data were analysed using reflexive thematic analysis (RTA), following Braun and Clarke’s approach [35–37]. The analysis (AW) was inductive and reflexive, with the researcher treated as an active interpreter rather than a neutral coding instrument.

All coding and theme development were undertaken by the first author and discussed with the second author (MB). The process involved repeated reading of transcripts, generation of initial codes, development and review of candidate themes, and refinement and definition of the final themes. Consistent with RTA, inter-rater reliability and coding consensus were not sought. Analytic decisions were documented through reflexive journalling and an audit trail. Recruitment was not based on a predetermined data-saturation criterion. Instead, sample adequacy was considered in relation to the richness and depth of participants’ accounts and their relevance to the aim [38,39].

### Reflexivity and trustworthiness

The first author (AW) is a registered Children’s and Young People’s Nurse and a higher education lecturer with experience of teaching in Further Education. At the time of the study, she was a full-time Master’s student in Public Health and health promotion. She has an interest in young people’s mental health and wellbeing and had no prior relationship with the participants. Her professional background will have shaped what she noticed and how she interpreted what she heard, and reflexive journalling allowed her to examine her assumptions and their potential influence on interpretation. Credibility was supported through repeated checking of transcripts against recordings, contextual engagement with the setting, and an audit trail.

## Findings

Twelve participants consented to take part; six were female, and six were male. Interviews lasted an average of 20 minutes (range 10–30). All twelve people recruited took part, and none withdrew. Participants described finding the farm through informal routes, including family or friends, internet searches, and employment support services. None had accessed it through a formal social prescribing referral pathway.

Four interconnected themes were developed (Table 1): (1) belonging in a supportive community; (2) finding purpose through meaningful participation; (3) personal growth and enhanced wellbeing; and (4) nature as the context for wellbeing. Social relationships, meaningful contribution, engagement with animals and the outdoor environment, and personal development were repeatedly described together rather than separately.

**Table 1.** Themes developed from interviews with young people at Swansea Community Farm.

| **Theme** | **What participants described** |
|---|---|
| 1. Belonging in a supportive community | A welcoming, inclusive and non-judgemental environment; reciprocal support from staff and peers; friendships and growing comfort interacting with others; a contrast with earlier educational, work and social settings. |
| 2. Finding purpose through meaningful participation | Structure, usefulness and a tangible contribution; responsibility for animals; learning practical skills and completing visible projects; a bridge towards work or further education. |
| 3. Personal growth and enhanced wellbeing | Gradual gains in mood, confidence, self-esteem, motivation and resilience; calm and respite from worries; support and coping rather than a cure; one account of friendship without a change in confidence. |
| 4. Nature as the context for wellbeing | Fresh air, open space and a change from home, school or college; animals as enjoyable and calming; physical work valued as part of meaningful work rather than as exercise; seasonal rhythms and a sense of contributing to something bigger. |

Themes were developed through reflexive thematic analysis of twelve individual interviews.

### Belonging in a supportive community

Participants described the farm as welcoming, inclusive and non-judgemental, often contrasting it with previous educational, employment or social environments in which they had felt isolated or misunderstood. Support was experienced as reciprocal rather than simply provided by staff. As P12 explained, *“Everyone’s quite helpful with each other. If something’s wrong, we’ll try and look after them or help them through whatever the issue is.”*

Relationships with staff and other participants were central. Several participants described developing friendships and becoming more comfortable interacting with others. P12 said, *“Interacting with other people has gotten better… before, I just didn’t really speak to other people. Here you’re speaking to people and interacting with them.”* Participants also valued the diversity of people at the farm and described a culture in which individuals could contribute according to their abilities.

### Finding purpose through meaningful participation

Participants consistently described taking part at the farm as giving them structure, purpose and opportunities to make a tangible contribution. Practical tasks included animal care, horticulture, maintenance, construction and conservation. Participants valued doing something useful, particularly compared with periods of inactivity at home. P12 described the value of *“getting to actually do something, get out of the house… and just having something to do with your hands.”*

Responsibility for animals was especially meaningful. Feeding animals, cleaning enclosures, monitoring welfare, and helping with animal husbandry were described as activities that generated trust and accountability. Participants also described satisfaction from learning practical skills and completing visible projects. Taking part could provide a bridge towards employment or further education by developing transferable skills and familiarity with workplace expectations.

### Personal growth and enhanced wellbeing

Participants described perceived improvements in mood, confidence, self-esteem, motivation and emotional resilience. Changes were generally described as gradual and cumulative. Several participants said they left the farm feeling happier or calmer and valued the opportunity to temporarily set aside everyday worries. P5 said, *“It makes me feel happier… when I’m here I don’t really think about everything else.”*

Confidence was linked to developing practical skills and interacting with other people in a supportive environment. Participants described becoming more willing to attempt unfamiliar tasks, taking on responsibility and, in some cases, informally supporting newer participants. Some linked their experiences to future aspirations in animal care, agriculture or environmental work.

Accounts were not uniformly positive. One participant valued the friendships made at the farm but did not perceive a meaningful improvement in confidence: *“Not really… I’ve made friends, but confidence? I don’t think so.”* Participants also acknowledged that taking part did not remove difficulties relating to education, employment, mental health or family circumstances.

They more often described it as a source of stability, coping and encouragement than as a cure.

### Nature as the context for wellbeing

The natural environment and animals were important parts of participants’ experience, but participants rarely described nature exposure alone as responsible for wellbeing. The farm was valued as an outdoor setting that felt different from home, school or college. Participants described fresh air, open space and a sense of being less constrained. They frequently described animals as enjoyable and calming, while animal care combined emotional connection with responsibility and purposeful activity.

Outdoor work also involved physical activity, but participants did not generally frame exercise as the main objective. Digging, lifting, walking, gardening and maintenance were experienced as enjoyable because they formed part of meaningful work. Seasonal rhythms, animal births, planting, harvesting and ongoing projects also gave participants a sense of contributing to something beyond themselves.

## Discussion

Mental health difficulties in young people are common, costly and tend to start early, yet we know little about how young people experience GSP or what makes it worth their time. We set out to explore how young people experienced taking part in a community farm programme in Wales and how they felt it influenced their mental health and wellbeing, so that what they value can inform the design and evaluation of GSP for their age group. We developed four interconnected themes: belonging in a supportive community; finding purpose through meaningful participation; personal growth and enhanced wellbeing; and nature as the context for wellbeing.

The main thing we take from these accounts is that the farm did not work through nature alone. What it appeared to do was bring three things together in one place: people who accepted participants and were glad to have them there; useful work that carried real responsibility and could be learned at their own pace; and time outdoors with animals and growing things. Participants rarely separated these. Belonging seemed to make it possible to try unfamiliar tasks. Responsibility and visible results built confidence and a sense of being useful. These are features of what is often called eudaimonic wellbeing, which is concerned with purpose, growth and flourishing rather than simply feeling good in the moment [40,41]. The outdoor setting and the animals gave the work its character and, for many, a welcome contrast with home, school or college. In terms of our objectives, participants had found the farm by informal routes (objective 1); they valued staff who treated them as individuals, peers who did not judge them and work that mattered (objective 2); they described gradual gains in mood, confidence and motivation rather than a cure (objective 3); and, taken together, their accounts suggest that GSP for young people may be best designed around belonging, meaningful roles and responsibility, with nature as the setting that makes these possible (objective 4).

### Belonging as the foundation

Belonging appeared to provide the foundation for much of what followed. Participants repeatedly emphasised being accepted, valued and supported, and described relationships with staff and peers as central. This closely mirrors what adults at the same farm have said. In a service-user study of the farm’s adult programme, participants described staff as welcoming and the farm as a kind of family, and a sense of belonging that in turn gave them purpose [21]. That young people and adults describe such similar processes suggests that these may be core features of the farm’s approach rather than something specific to age. They also resonate with the few studies of young people in GSP-type provision: young care leavers in a socially prescribed gardening group described social belonging and a safe space [28], and young pregnant women valued nature-based activities partly for the social support they offered [27].

This also complements the wider GSP literature, which considers both environmental and social processes [15,16,31].

Self-determination theory offers one way to read these accounts. It proposes that wellbeing depends on feeling related to others, competent and autonomous [42], and participants’ descriptions of acceptance (relatedness) and of mastering tasks and skills (competence) fit that picture. Others have used the same theory to understand adult social prescribing and found that trusting relationships with staff and peers and the chance to develop new skills were central [23]. Social identity may be a further route, since being part of a group with a shared purpose is thought to be one way social prescribing builds connection [22]. Seen through an ecological lens [43], the farm gave young people a setting in which relationships, roles and the physical environment reinforced one another, which may be why participants found them hard to pull apart.

### Meaningful participation and what counts as an outcome

Meaningful participation was equally prominent. Participants valued practical work, responsibility, learning and visible contribution, and described these in terms of usefulness, purpose and competence rather than symptom reduction. Similar themes appear in adult accounts of this scheme, where skills and training created ‘ripple effects’ that reached into life beyond the farm [21]. Ripple effects of nature-based activity beyond the session itself have also been described in a surf-therapy programme for adults with acquired brain injury [44]. They also sit alongside wider evidence that volunteering is associated with lower depression and higher life satisfaction and wellbeing, although that evidence comes mainly from adults [45]. Some participants linked their time at the farm to future aspirations in animal care, agriculture or environmental work, which may matter particularly for young people at a stage when disengagement from education and work is a real risk [6].

This has a direct implication for evaluation. Conventional symptom measures may not capture belonging, confidence, purpose and meaningful contribution, yet these were the outcomes participants talked about [19,21]. Evaluations of GSP for young people that rely only on symptom scales risk missing what young people value and, equally, what may be driving any change. Measures of belonging, self-efficacy and purpose, developed with young people, would be a sensible addition. Participants were also clear that the farm was not a cure. They described stability, coping and encouragement, and one participant did not notice a change in confidence at all. That fits with GSP as a complement to, rather than a replacement for, clinical care.

### Volunteering or green social prescribing?

Because participants were volunteers, a reader may reasonably ask whether this is a study of volunteering rather than GSP. In the findings, we have written about taking part at the farm rather than about volunteering, to keep the focus on the experience rather than the role. But the overlap is real. GSP schemes often take the form of practical gardening, conservation or farm tasks [8,9], and the farm’s volunteering is the vehicle through which its mental health and wellbeing aims are pursued. The features young people valued, such as being needed, having responsibility and contributing to something visible, are also features commonly attributed to volunteering [45]. We therefore see volunteering less as a competing explanation than as one of the active ingredients of this kind of provision. It also raises a design question. A role that asks for responsibility and reliability may suit some young people better than others, and social prescribers in South Wales have described many of their clients as not yet ready for outdoor group activity [20]. Those who most need support may be the least ready to take it up.

### Nature as an enabling context

These findings caution against treating ‘nature-based’ as synonymous with passive exposure to nature. Participants were actively caring for animals, gardening, maintaining the farm and working alongside other people, and nature appeared to function as an enabling context for occupation and relationships rather than as the active ingredient in itself. Adults at the same farm valued nature exposure and connection but placed them within a supportive community [21], and a qualitative study of a campus community garden found that connectedness to nature, growing food and community building were valued together [29]. Staff in child and adolescent mental health services have also suggested that shared outdoor activity can take the pressure off conversation for young people [5]. Participants did not describe it in these terms, but working side by side on shared tasks may have offered a lower-pressure way to build relationships than formal conversation, which would fit the importance they gave to belonging.

Our interpretation is compatible with psychological theories of restorative environments, in which natural settings support recovery from stress and mental fatigue [46,47], and the calm, fresh air and open space that participants described are in keeping with those ideas. However, greenspace exposure and connection to nature have been linked to health outcomes mainly in observational work [48,49], and our study did not measure nature connectedness or establish a mechanism of change.

Animal care was particularly notable. Participants described animals as enjoyable, calming and emotionally rewarding, and caring for them brought responsibility and practical competence. Farm animals have been described as important to many care-farm participants, including young people with behavioural difficulties [50], yet human–animal interaction remains less prominent in the wider GSP evidence base. It may be a distinctive feature of community farms compared with other GSP settings and deserves attention in both design and research.

### How young people reached the farm: self-referral and what counts as social prescribing

None of the participants had entered the farm through a formal GSP referral pathway. They had found it through family or friends, internet searches and employment support services. This does not establish that formal referral pathways are ineffective. Rather, it indicates that these young people reached a community asset through informal routes and experienced it as relevant to their wellbeing. It matters because whether self-referral ‘counts’ as social prescribing is contested. The Welsh national framework recognises self-referral as one route [10], but in a previous stakeholder study in South Wales people disagreed about whether schemes entered without a link worker or health professional should be called social prescribing at all [20]. Our findings add the perspective of young people who arrived this way: they found a place they experienced as supportive, and they did so without being referred.

At the same time, relying on informal discovery carries risks. The routes participants described depend on having family or friends who know about the farm, on being in contact with employment services, or on knowing what to search for. Young people without those connections may never find provision of this kind. The formal route may not currently fill the gap, because providers in South Wales reported receiving fewer referrals into nature-based schemes than they had anticipated [20]. In practice, this points towards offering several routes at once: self-referral that is easy for young people to find and use, referral from employment and education services where young people already are, and link-worker referral for those who need support to take a first step. Whether formal referral changes engagement or outcomes is a question for future research.

### Equity and access

Community-based provision can potentially extend support beyond clinical services, but transport, awareness, socioeconomic circumstances and other barriers shape who takes part [51,52]. Provision is also fragile. Third sector providers in South Wales have described dependence on short-term grant funding [20], and a mixed-methods study of GSP for young pregnant women reported insufficient funding and disappointing experiences with some programmes [27]. Relational continuity with staff appeared important in the adult study of this farm [21], so unstable funding may matter particularly for young people who need time to build trust. We did not recruit to examine inequality and cannot say how accessible the farm is to young people facing the greatest barriers.

### Implications for practice, policy and evaluation

Taken together, these findings suggest four things for those designing GSP for young people. First, provision may work best when it is built around belonging, meaningful roles and real responsibility, delivered by staff who treat young people as individuals, with nature as the setting rather than the whole intervention. Second, ‘nature-based’ should not be read as passive exposure, and schemes that offer little opportunity to contribute or to work alongside others may not offer the same experience. Third, young people need routes in that they can find and use themselves as well as through referral, and these should be visible in the places they already go, including online and through employment support. Fourth, evaluation should include outcomes such as belonging, confidence and purpose, ideally developed with young people, alongside symptom measures.

## Limitations

The findings come from a single community farm and twelve participants. The study aimed for depth rather than generalisation, so readers will need to judge how far the accounts apply to other settings with similar features. Reflexive thematic analysis does not rely on a fixed sample size or on saturation; the question is whether the data are rich enough to support the interpretation offered [38,39], and qualitative studies of comparable scope exist, such as the study of six young care leavers in a community gardening group [28]. Studies at further sites, including rural and larger-scale provision, would clarify which features are transferable to other settings.

Participants were current attendees recruited through the farm manager. We therefore heard from young people who had stayed, who felt comfortable enough to take part, and who may have been inclined to speak well of a place they valued. We did not hear from young people who left, never settled, or could not access the farm, and their experiences may differ. There was no comparison group, and we did not measure mental health outcomes, so perceived improvements cannot be interpreted as evidence of causal effectiveness.

Participants were not formally referred through a GSP pathway, so the study cannot consider referral processes or link-worker support, and the findings may not apply to young people who arrive by referral, who may differ in needs, readiness and expectations. We have described the farm as a form of GSP because the national framework recognises self-referral [10], but that framing is not universally accepted [20]. The study was also not designed to examine inequality, and we collected no data on socioeconomic circumstances, transport or other barriers. national framework recognises self-referral [10], but that framing is not universally accepted [20]. The study was also not designed to examine inequality, and we collected no data on socioeconomic circumstances, transport or other barriers.

Transcripts and findings were not returned to participants. In reflexive thematic analysis, themes are the researcher’s interpretation of patterns across the dataset rather than a verifiable account of what any one person said, so member checking is not treated as a test of validity [35,53], although member reflections might have enriched the analysis [53]. Coding was undertaken by one researcher (AW) and discussed with the second author. Reflexive journalling and an audit trail were used to examine how her nursing and teaching background shaped interpretation, but a different analyst might have developed different themes [35,36].

## Future directions

Several questions follow from this study. How do young people discover and access provision like this, and does formal referral change engagement or outcomes? Studies that compare young people who arrive by self-referral, link-worker referral and other statutory routes would help answer this. What happens over time: are the gains participants described sustained, and what happens when young people leave? Longitudinal work and interviews with young people who disengaged would also address the limits of a sample made up of current attenders. Research at more sites, in rural and urban settings and with more diverse groups of young people, would show which features of community farms and similar provision are transferable. Finally, building on the framework for the ‘active ingredients’ of nature-based social prescribing that is being developed from adult accounts [21], future work could co-develop with young people outcome measures that capture belonging, confidence and purpose.

## Conclusions

Young people in this study described community farm participation as supporting their mental health and wellbeing through interconnected experiences of belonging, meaningful participation, personal growth and engagement with nature and animals. Nature was valued as part of a wider social and occupational environment rather than as an isolated therapeutic exposure. For GSP to work for young people, provision may need to prioritise belonging, meaningful roles and responsibility, offer routes into provision that young people can find and use themselves as well as through referral, and be evaluated against outcomes that matter to them. Further research should examine whether these processes are sustained over time, how young people access community nature-based provision, and whether similar processes are present in other settings.

## Data Availability

The minimal anonymised data is available on request from the corresponding author and will be provided if in line with the ethical consent guidelines the study was approved under.

## Acknowledgments

The authors thank Swansea Community Farm and its staff for facilitating access to the setting and participants, and the young people who shared their experiences.

## Funding

The author(s) received no specific funding for this work.

## Competing interests

The authors have declared that no competing interests exist.

## Supporting information

**S1.** Checklist. COREQ checklist. Consolidated criteria for reporting qualitative research (COREQ) 32-item checklist, completed for this study.

## Notes

### Competing Interest Statement

The authors have declared no competing interest.

### Author Declarations

This study was approved by the Faculty of Medicine, Health and Life Sciences, School of Health and Social Care Research Ethics Committee Swansea University on 5.5.26 ref number # 2 2026 15441 16277

## References

1. World Health Organization. Adolescent mental health. Geneva: WHO; 2021. Available from: https://www.who.int/news-room/fact-sheets/detail/adolescent-mental-health

2. Sawyer SM, Azzopardi PS, Wickremarathne D, Patton GC. The age of adolescence. Lancet Child Adolesc Health. 2018;2(3):223–228. doi: 10.1016/S2352-4642(18)30022-1

3. Solmi M, Radua J, Olivola M, Croce E, Soardo L, Salazar de Pablo G, et al. Age at onset of mental disorders worldwide: large-scale meta-analysis of 192 epidemiological studies. Mol Psychiatry. 2022;27(1):281–295. doi: 10.1038/s41380-021-01161-7

4. NHS Digital. Mental health of children and young people in England, 2022 - wave 3 follow-up to the 2017 survey. Leeds: NHS Digital; 2022.

5. Chapman B, Mitchell SB, Hardwick R, Bonnici H, Banks H, Mareva S, et al. A qualitative exploration of the potential benefits of nature-based approaches for staff and children and young people in child and adolescent mental health services. PLOS Ment Health. 2025;2(7):e0000389. doi: 10.1371/journal.pmen.0000389

6. Gariépy G, Danna SM, Hawke L, Henderson J, Iyer SN. The mental health of young people who are not in education, employment, or training: a systematic review and meta-analysis. Soc Psychiatry Psychiatr Epidemiol. 2022;57(6):1107–1121. doi: 10.1007/s00127-021-02212-8

7. Cardoso F, McHayle Z. The economic and social costs of mental ill health. London: Centre for Mental Health; 2024. Available from: https://www.centreformentalhealth.org.uk/publications/the-economic-and-social-costs-of-mental-ill-health/

8. Polley M, Fleming J, Anfilogoff T, Carpenter A. Making sense of social prescribing. London: University of Westminster; 2017.

9. Husk K, Lovell R, Garside R, Cooper C. Social prescribing: where is the evidence? Br J Gen Pract. 2019;69(678):6–7.

10. Welsh Government. National framework for social prescribing. Cardiff: Welsh Government; 2024. Available from: https://www.gov.wales/national-framework-social-prescribing

11. Well-being of Future Generations (Wales) Act 2015. anaw 2.

12. Welsh Government. A healthier Wales: our plan for health and social care. Cardiff: Welsh Government; 2018.

13. Welsh Government. Mental health and wellbeing strategy 2025 to 2035. Cardiff: Welsh Government; 2025.

14. Welsh Government. Core standards and values for social prescribing in Wales. Cardiff: Welsh Government; 2026.

15. Thomas T, Aggar C, Baker J, Massey D, Thomas M, D’Appio D, et al. Social prescribing of nature therapy for adults with mental illness living in the community: a scoping review of peer-reviewed international evidence. Front Psychol. 2022;13:1041675. doi: 10.3389/fpsyg.2022.1041675

16. Menhas R, Yang L, Saqib ZA, Younas M, Saeed MM. Does nature-based social prescription improve mental health outcomes? A systematic review and meta-analysis. Front Public Health. 2024;12:1228271. doi: 10.3389/fpubh.2024.1228271

17. Sachs AL, Kolster A, Wrigley J, Papon V, Opacin N, Hill N, et al. Connecting through nature: a systematic review of the effectiveness of nature-based social prescribing practices to combat loneliness. Landsc Urban Plan. 2024;248:105071.

18. Leavell MA, Leiferman JA, Gascon M, Braddick F, Gonzalez JC, Litt JS. Nature-based social prescribing in urban settings to improve social connectedness and mental well-being: a review. Curr Environ Health Rep. 2019;6(4):297–308. doi: 10.1007/s40572-019-00251-7

19. Ashe MC, dos Santos IK, Alfares H, Chudyk AM, Esfandiari E. Outcomes and instruments used in social prescribing: a modified umbrella review. Health Promot Chronic Dis Prev Can. 2024;44(6):244–269.

20. Brown M, Aylett KS. Interrogating green social prescribing in South Wales; a multi-stakeholder qualitative exploration. PLoS One. 2025;20(1):e0314107. doi: 10.1371/journal.pone.0314107

21. Brown M, Jenkins C, Froggatt AL. A qualitative exploration of a UK, nature-based social prescribing scheme; the service user perspective. Soc Med. 2026;19(2):164–173. doi: 10.71164/socialmedicine.v19i2.2026.2255

22. Staras CO, Wakefield JRH, McDermott DT, Jones BA. An evaluation of the role of social identity processes for enhancing health outcomes within UK-based social prescribing initiatives designed to increase social connection and reduce loneliness: a systematic review. J Community Appl Soc Psychol. 2024;34(5):e2878.

23. Bhatti S, Rayner J, Pinto AD, Mulligan K, Cole DC. Using self-determination theory to understand the social prescribing process: a qualitative study. BJGP Open. 2021;5(2).

24. Olsson A, Stapley E, Russell A, Hayes D. Young people’s views of social prescribing as an approach to improving mental health: a qualitative study. Health Soc Care Community. 2025;2025:2969967. doi: 10.1155/hsc/2969967

25. Tillmann S, Tobin D, Avison W, Gilliland J. Mental health benefits of interactions with nature in children and teenagers: a systematic review. J Epidemiol Community Health. 2018;72(10):958–966. doi: 10.1136/jech-2018-210436

26. Overbey TA, Diekmann F, Lekies KS. Nature-based interventions for vulnerable youth: a scoping review. Int J Environ Health Res. 2023;33(1):15–53.

27. Sands G, Blake H, Carter T, Spiby H. Nature-based interventions in the UK: a mixed methods study exploring green prescribing for promoting the mental wellbeing of young pregnant women. Int J Environ Res Public Health. 2023;20(20):6921.

28. Moore EJ, Thew M. Exploring the perspectives of ’young adults’ (18-24) who have been in formal care and their experiences of attending a socially prescribed community allotment gardening group. Br J Occup Ther. 2023;86(1):26–32. doi: 10.1177/03080226221117447

29. D’Andrea Brooks O, Thanganathan R, Gittings L. Growing third places: a qualitative study of experiences and perceived benefits of a campus community garden as a nature-based health intervention. PLoS One. 2025;20(12):e0338602. doi: 10.1371/journal.pone.0338602

30. Murray J, Wickramasekera N, Elings M, Bragg R, Brennan C, Richardson Z, et al. The impact of care farms on quality of life, depression and anxiety among different population groups: a systematic review. Campbell Syst Rev. 2019;15(4):e1061.

31. Bragg R, Atkins G. A review of nature-based interventions for mental health care. Natural England; 2016.

32. Creswell JW, Poth CN. Qualitative inquiry and research design. 4th ed. Thousand Oaks, CA: SAGE; 2018.

33. Denzin NK, Lincoln YS, editors. The SAGE handbook of qualitative research. 5th ed. Thousand Oaks, CA: SAGE; 2018.

34. Tong A, Sainsbury P, Craig J. Consolidated criteria for reporting qualitative research (COREQ): a 32-item checklist for interviews and focus groups. Int J Qual Health Care. 2007;19(6):349–357. doi: 10.1093/intqhc/mzm042

35. Braun V, Clarke V. Thematic analysis: a practical guide. London: SAGE; 2021.

36. Braun V, Clarke V. Conceptual and design thinking for thematic analysis. Qual Psychol. 2022;9(1):3–26.

37. Braun V, Clarke V. Toward good practice in thematic analysis. Int J Qual Methods. 2023;22:1–16.

38. Malterud K, Siersma VD, Guassora AD. Sample size in qualitative interview studies: guided by information power. Qual Health Res. 2016;26(13):1753–1760. doi: 10.1177/1049732315617444

39. Braun V, Clarke V. To saturate or not to saturate? Questioning data saturation as a useful concept for thematic analysis and sample-size rationales. Qual Res Sport Exerc Health. 2021;13(2):201–216. doi: 10.1080/2159676X.2019.1704846

40. Ryan RM, Deci EL. On happiness and human potentials: a review of research on hedonic and eudaimonic well-being. Annu Rev Psychol. 2001;52:141–166.

41. Kemp AH, Fisher Z. Wellbeing, whole health and societal transformation: theoretical insights and practical applications. Glob Adv Health Med. 2022;11:21649561211073076.

42. Ryan RM, Deci EL. Self-determination theory: basic psychological needs in motivation, development, and wellness. New York: Guilford Press; 2017.

43. Bronfenbrenner U. The ecology of human development. Cambridge, MA: Harvard University Press; 1979.

44. Wilkie L, Fisher Z, Kemp AH. The ‘rippling’ waves of wellbeing: a mixed methods evaluation of a surf-therapy intervention on patients with acquired brain injury. Sustainability. 2022;14(15):9605. doi: 10.3390/su14159605

45. Jenkinson CE, Dickens AP, Jones K, Thompson-Coon J, Taylor RS, Rogers M, et al. Is volunteering a public health intervention? A systematic review and meta-analysis of the health and survival of volunteers. BMC Public Health. 2013;13:773. doi: 10.1186/1471-2458-13-773

46. Kaplan R, Kaplan S. The experience of nature. Cambridge: Cambridge University Press; 1989.

47. Ulrich RS, Simons RF, Losito BD, Fiorito E, Miles MA, Zelson M. Stress recovery during exposure to natural and urban environments. J Environ Psychol. 1991;11(3):201–230.

48. Twohig-Bennett C, Jones A. The health benefits of the great outdoors. Environ Res. 2018;166:628–637.

49. Kenyon A, Chauhan T, Reynolds K, Pywell S, Shaikh R, Ajiboye A. Connection to nature in nature-based social prescribing. Soc Sci Med. 2026;397:119133.

50. Hassink J, De Bruin SR, Berget B, Elings M. Exploring the role of farm animals in providing care at care farms. Animals (Basel). 2017;7(6):45. doi: 10.3390/ani7060045

51. Marmot M, Allen J, Boyce T, Goldblatt P, Morrison J. Fair society, healthy lives. London: Institute of Health Equity; 2010.

52. Venter ZS, Barton DN, Gundersen V, Figari H, Nowell M. Urban nature and health equity. Landsc Urban Plan. 2023;232:104684.

53. Smith B, McGannon KR. Developing rigor in qualitative research: problems and opportunities within sport and exercise psychology. Int Rev Sport Exerc Psychol. 2018;11(1):101–121. doi: 10.1080/1750984X.2017.1317357

